# Association of Network Connectivity via Resting State Functional MRI with Consciousness, Mortality, and Outcomes in Neonatal Acute Brain Injury

**DOI:** 10.1101/2021.10.18.21265147

**Authors:** Varina L. Boerwinkle, Bethany Sussman, Iliana Manjón, Lucia Mirea, Saher Suleman, Sarah N. Wyckoff, Alexandra Bonnell, Andrew Orgill, Deborah Tom

**Affiliations:** Division of Pediatric Neurology, Barrow Neurological Institute at Phoenix Children’s Hospital, 1919 E. Thomas Rd, Phoenix, AZ 85016 USA; Department of Neuroscience Research, Barrow Neurological Institute at Phoenix Children’s Hospital, 1919 E. Thomas Rd, Phoenix, AZ 85016 USA; Department of Clinical Research, Phoenix Children’s Hospital, 1919 E. Thomas Rd, Phoenix, AZ 85016 USA; Division of Neonatology, Phoenix Children’s Hospital, 1919 E. Thomas Rd, Phoenix, AZ 85016 USA; University of Arizona College of Medicine – Tucson, 1501 N. Campbell Ave, Tucson, AZ, 85724 USA

**Keywords:** Resting state functional MRI (rs-fMRI), neonatal acute brain injury (ABI), hypoxic ischemic encephalopathy (HIE), connectivity, consciousness

## Abstract

**Background:** An accurate and comprehensive test of integrated brain network function is needed for neonates during the acute brain injury period to inform on morbidity. This retrospective cohort study aimed to assess whether integrated brain network function by resting state functional MRI, acquired during the acute period in neonates with brain injury, is associated with acute exam, neonatal mortality, and 5-month outcomes.

**Methods:** This study included 40 consecutive neonates with resting state functional MRI acquired 1-22 days after suspected brain insult from March 2018 to July 2019 at Phoenix Childrens Hospital. Acute period exam and test results were assigned ordinal scores based on severity as documented by respective treating specialists. Analyses (Fisher exact, Wilcoxon-rank sum test, ordinal/multinomial logistic regression) examined association of resting state networks with demographics, presentation, neurological exam, electroencephalogram, anatomical MRI, magnetic resonance spectroscopy, passive task functional MRI, and outcomes of discharge condition, outpatient development, motor tone, seizure, and mortality.

**Results:** Subjects had a mean (standard deviation) gestational age of 37.8 (2.6) weeks, a majority were male (63%), with diagnosis of hypoxic ischemic encephalopathy (68%). Other findings at birth included mild distress (48%), moderately abnormal neurological exam (33%), and consciousness characterized as awake but irritable (40%). Significant associations after multiple testing corrections were detected for resting state networks: basal ganglia with outpatient developmental delay (odds ratio [OR], 14.5; 99.4% confidence interval [CI], 2.00-105; *P*<.001) and motor tone/weakness (OR, 9.98; 99.4% CI, 1.72-57.9; *P*<.001); language/frontal-parietal network with discharge condition (OR, 5.13; 99.4% CI, 1.22-21.5; *P*=.002) and outpatient developmental delay (OR, 4.77; 99.4% CI, 1.21-18.7; *P=*.002); default mode network with discharge condition (OR, 3.72; 99.4% CI, 1.01-13.78; *P=*.006) and neurological exam (*P=*.002 (FE); OR, 11.8; 99.4% CI, 0.73-191; *P=*.01 (OLR)); seizure onset zone with motor tone/weakness (OR, 3.31; 99.4% CI, 1.08-10.1; *P=*.003). Resting state networks were not detected in only three neonates, who died prior to discharge.

**Conclusions:** This study provides level 3 evidence (OCEBM Levels of Evidence Working Group) that the degree of abnormality of resting state networks in neonatal acute brain injury is associated with acute exam and outcomes. Total lack of brain network detection was only found in patients who did not survive.

## 1. Introduction

In neonatal acute brain injury (ABI), exam and test markers of integrated brain network function (IBNF) are needed to characterize disorders of consciousness (DoC), recovery potential, and inform withdrawal of life sustaining therapy determinations (WLST), with profound consequences (Pant, 2021; Kondziella et al., 2020; Michel et al., 2019; Douglas-Escobar and Weiss, 2015; Downar et al., 2016; Demertzi et al., 2011; Turgeon et al., 2011). Confounds such as complex movements which may be intentional, seizure, reflexes, or automatisms, with varying treatments and outcomes, (Han et al., 2006; Saposnik et al., 2005) contribute to DoC misdiagnosis in 46% of adults (Schnakers et al., 2009). Higher missed-or-no DoC diagnosis in neonates due to less-informed DoC DoC definitions, validated exam markers, and diagnostics (Giacino et al., 2018; Nakagawa et al., 2012; Bruno et al., 2011; Schnakers et al., 2009; Machado, 2008) likely reduces precision in diagnosis-informed treatments, outcome predictions, and WLST determinations (Pant, 2021; Downar et al., 2016; Turgeon et al., 2011; Devictor et al., 2008).

IBNF is pivotal in adult DoC diagnosis and recovery prediction (Hahn et al., 2021; Michel et al., 2019; Giacino et al., 2018; Sair et al., 2018; Kondziella et al., 2017; Falletta Caravasso et al., 2016; Rosazza et al., 2016; Silva et al., 2015). IBNF modalities include task (stimulation)-based and resting state electroencephalogram (EEG) and functional magnetic resonance imaging (fMRI), wherein the measure indicative of the highest capacity for consciousness ***supplants*** other modalities including exam in adults with subacute to chronic DoC (Kondziella et al., 2020). However, such diagnostic precision need is greater in acute than chronic brain injury given the higher mortality and rate of WLST in the acute period, which likely also effects pediatric and neonatal patients.

Of the DoC-guideline IBNF modalities available, much progress has been made in neonates with resting state functional MRI (RS). In *heathy* neonates, resting state networks (RSN) are detectable by the 3^rd^ trimester (Doria et al., 2010). The default mode (DMN), attention, and frontal-parietal (FP) RSN are detectable by term (van den Heuvel et al., 2015; Fransson et al., 2009). In neonates with *mild to mild-moderate* ABI, RSN metrics are predictive of neurodevelopmental outcomes (Ferradal et al., 2018; He et al., 2018; Mak et al., 2018; Shi et al., 2018; Doria et al., 2010; Smith et al., 2009; Damoiseaux et al., 2006; Jenkinson et al., 2002). Prematurity affects long-range and frontal connectivity, (Della Rosa et al., 2021; Bouyssi-Kobar et al., 2019; Strahle et al., 2019; He et al., 2018; Linke et al., 2018; Wheelock et al., 2018; Rogers et al., 2017; Smyser et al., 2010) and hypoxic ischemic encephalopathy (HIE) (Li et al., 2019) affects the language, vision, and sensory-motor networks. In acute *mild to moderate* ABI, RS is also predictive of neurodevelopmental outcomes (Della Rosa et al., 2021; Bouyssi-Kobar et al., 2019; Li et al., 2019; Strahle et al., 2019; He et al., 2018; Linke et al., 2018; Wheelock et al., 2018; Rogers et al., 2017; Smyser et al., 2010). However, only one large neonatal-ABI-RS study performed on a 1.5 Tesla MRI, which has significantly lower fMRI signal detection capacity than higher Tesla MRI (Krasnow et al., 2003) included ***<u>severe</u>*** ABI: the population most likely to have WLST determinations in the acute period made, and anatomical MRI (a-MRI), exam, and brain-behavior evaluation. It showed RS-motor outcome association, ***<u>irrespective of pathological grouping</u>***, with demographics, presentation, course severity clinical factors, and a-MRI severity, while relationships between RSN, mortality, acute exam, and a-MRI were not determined (Bhroin et al., 2021; Linke et al., 2018). Lastly, RSN alterations from perinatal brain injury are detectable years later and correspond to developmental outcomes, indicating the RS findings from neonatal ABI are durable and meaningful (Bhroin et al., 2021). Thus, the gap in utility of RS in the full spectrum of ABI severity against outcomes beyond motor is unknown, and validation of the original motor finding is needed.

Importantly, developing biomarkers of network pathology in heterogeneous cohorts of brain injury are of high priority, according to recent expert opinion consensus (Edlow et al., 2020; Kondziella et al., 2020; LaRovere and Tasker, 2020; Provencio et al., 2020). The expert consensus reasoning is such that the effect on IBNF on the brain insult, rather than the particular insult subtype, is thought to be the most likely determining factor in capacity for recovery. Thus, narrowed injury causation subtype designed trial is thought to be less favorable in demonstrating ultimate clinical utility.

Exploring connectivity relationship to the controversial consciousness exam in neonates is needed. Without supratentorial network activity confirmation, such as EEG sleep-wake brain state changes, neonatal consciousness behaviors have been proposed by some to be brainstem-driven, with no link to recovery potential (Ashwal et al., 1990). Alternatively, in non-anencephalic, stimulus-brainstem-driven neonatal behaviors may still predict future consciousness (Ishaque et al., 2017; Del Giudice et al., 2016). Supportively, lack of arousal behavior is reliable enough, by current expert consensus, to be a key component of the brain death (BD) determination in neonates (Nakagawa et al., 2012; Nakagawa et al., 2011).

Thus, this study aims to evaluate RS in *mild to severe* neonatal ABI with acute period neurological/consciousness exam, mortality, other tests, and outcomes.

## 2. Methods

Study patients included consecutive ABI neonates during March 2018 to July 2019, with clinical RS and MRI evaluated as standard of care. The institutional review board approved this retrospective study with waived consent. Data were collected from the medical records including demographics (sex, gestational age), diagnosis, presentation (Sarnat-diagnosis/mental status (Mrelashvili et al., 2020)), and exams/tests (neurological exam, consciousness, anatomical MRI (a-MRI), task-fMRI, RS results, magnetic resonance spectroscopy (MRS), EEG) during the acute period, discharge condition, mortality, and outpatient outcomes (development, motor tone, strength, seizure). If the element’s categorization exact wording was not specifically documented within the treating physician’s acute period charted note, then two separately blinded study personnel made the determination based on the wording available, with plan for third research personnel if disagreement occurred; however, none occurred. The number of patients receiving each assessment is shown in **Table 1**. The acute-period HIE severity, diagnoses, presentation, and neurological and consciousness exams were documented prior to MRI with RS scan. Initial EEG reads occurred prior to MRI scan, and after HIE and presentation severity determination but before final diagnoses determinations.

**Table 1.**
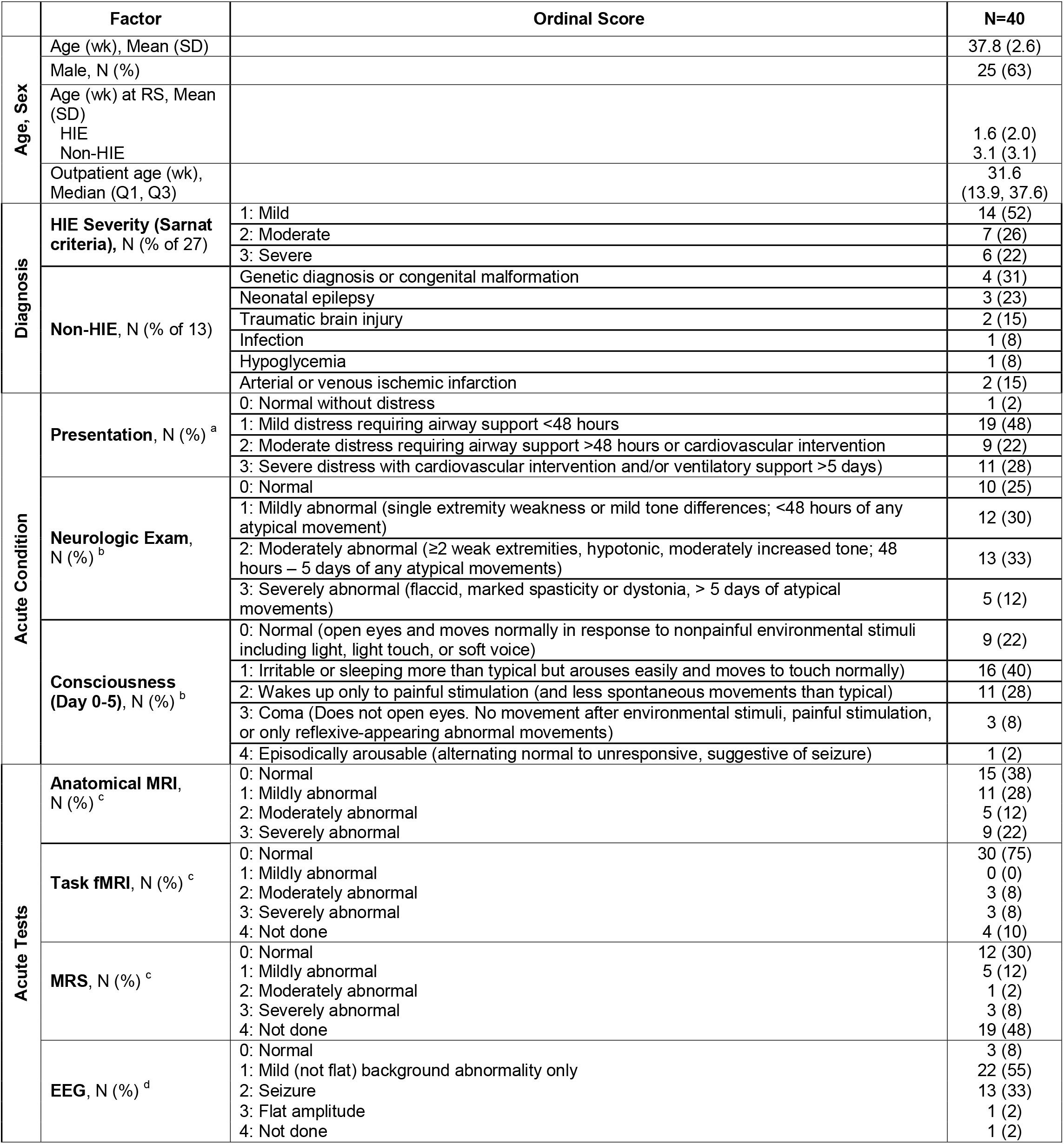
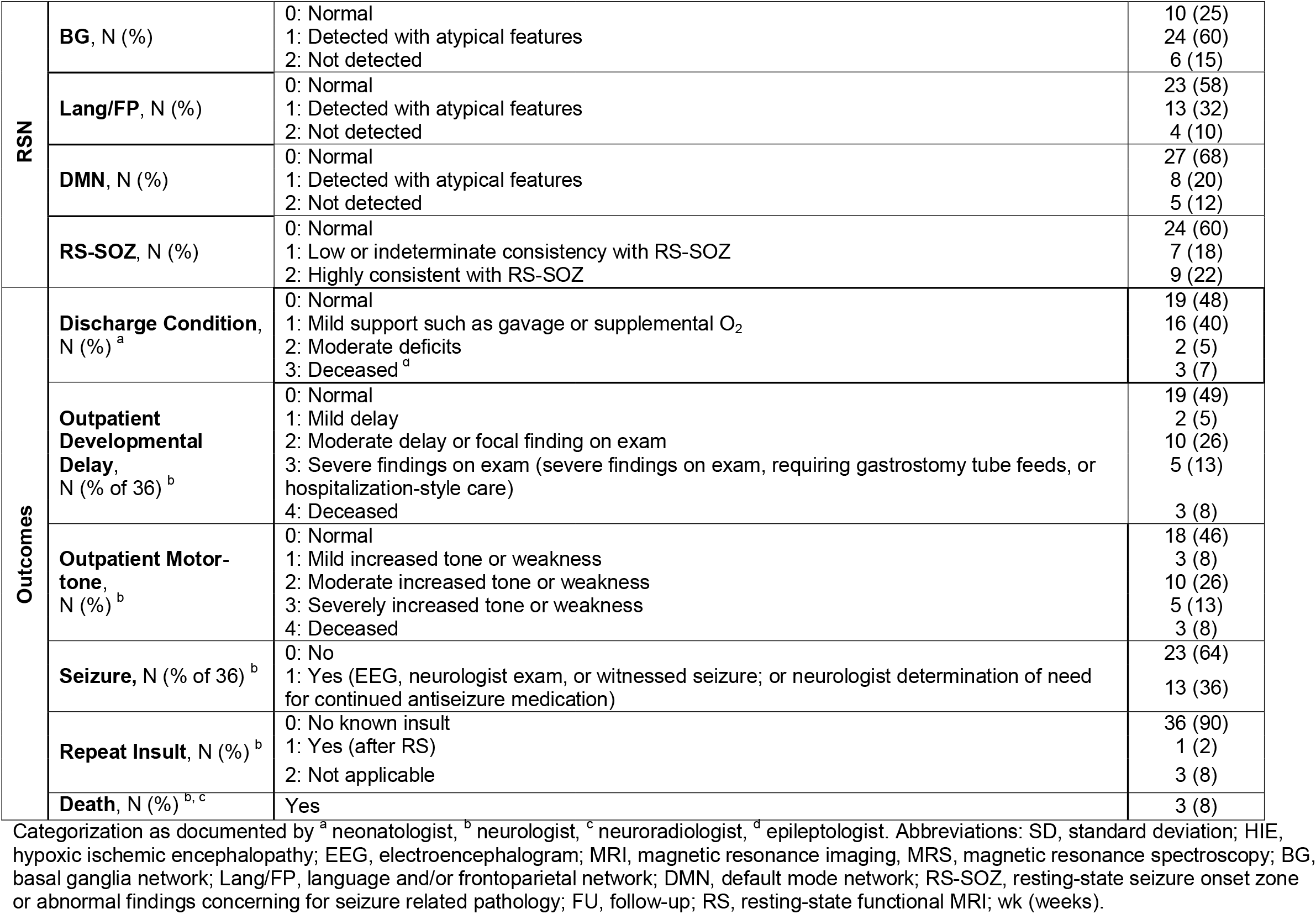
Demographics and Ordinal Scores of Acute Clinical Findings Tests and Outcomes

The acute-period and outcome diagnosis and exam findings were categorized as ordinal scores based on factors documented by the respective treating pediatric specialists further described in the following sections 2.1 through 2.3.

### 2.1 Acute-Period Diagnoses

<u>HIE</u> was determined by Sarnat-criteria (Mrelashvili et al., 2020), as documented by the neonatologist. <u>HIE Severity and non-HIE diagnoses</u> were determined by the discharging neonatologists final report.

<u>Presentation severity</u>/distress categorization was determined by the treating neonatologist’s determination for need of airway or ventilatory support, and cardiovascular intervention, and the respective interventions length of implementation, as detailed in **Table 1**.

### 2.2 Neurological and Consciousness Exams

<u>Neurological and consciousness exams</u> were determined by the initial consulting neurocritical care provider’s exam, who are pediatric neurologists, on the day of consult, which is typically on the day of admission (or the following day if admitted overnight). Repeat exams were documented if new additional events occurred. These may include new weakness, worsened encephalopathy, movements concerning for seizure, or new abnormal findings on the EEG. Scales are lacking for neonatal acute period factors and outcomes that are validated in a mixed pathology and full spectrum severity peri-term gestation population, or any neonatal population related to consciousness, other than those denoted. Thus, these elements were categorized by ordinal scales designed to capture the severity range based on the neurologist’s documentation of weakness, tone difference, abnormal movements, lack of normal movement, and length of time of ongoing difference from normal, as delineated in **Table 1**.

#### Repeat insult

To differentiate outcomes related to the acute period exams, differentiation between those with a second separate brain insult were recorded. Those who died prior to discharge were separately noted.

#### Death

Only those who died prior to discharge were recorded. No further deaths occurred at time of last available study follow-up (within 6 months).

### 2.3 Outcome Diagnoses and Exams

<u>Discharge condition</u> was determined by the discharging neonatologist according to their documentation of ongoing need for gavage feeding or supplemental oxygen being the mild category, and all other ongoing support or detectable abnormalities being categorized as “moderate deficit”. The most severe category included those who did not survive.

<u>Outpatient development, motor-tone, and seizure</u> were determined by the pediatric neurologist through the last available follow-up (within 6 months) through authorized study collection end (**Table 1**). Developmental delays diagnosed in the medical record and any ongoing focal neurological deficits factored into the follow-up “outpatient development” categorization. Severe findings on exam included ongoing need for gastrostomy tube feeds, airway clearance by deep suctioning, or ventilatory support. The “motor-tone” outcome is an abbreviated combination of motor abnormalities including both weakness and tone. The severity rating was based entirely on the documenting neurologist’s impression of degree of difference from normal (**Table 1**). The “seizure” outcome denotes an ongoing concern for either a possible seizure occurring after discharge, either by EEG-capture, neurologists’ exam, or caretaker witnessed event *or* treating neurologist’s impression of need for ongoing anti-seizure medication.

### 2.4 Acute Period MRI and EEG

#### 2.4.1 ICU MRI safety, anesthesia, and scan timing

To receive a clinical MRI, all patients were determined to be safe and stable for transport to the scanner by the neonatologist and received continuous observation from the respective hospital care teams. Total scan time was approximately one hour. No safety event occurred during or related to scanning. As this is a retrospective study, no study care modifications occurred. Patients at the time of scan were not receiving anesthetic agents, as per their care plans. To minimize movement during imaging, infants were fed and swaddled, with multiple levels of ear protection, including earplugs, earmuffs, and standard headphones. Awake infants were rescheduled for later scan times as needed. As such, neonates with HIE, also diagnosed by their treating neonatologist (Jacobs et al., 2011; Azzopardi et al., 2009) received 48 hours of therapeutic hypothermia and medications for symptomatic relief of cold temperature. After the end of cooling, clinical MRI with RS was performed on day 3-5. Infants with non-HIE brain MRI indications, received MRI with RS at more variable times according to clinical indications within the first month of life. Image and EEG severity categorization were determined by the interpreting pediatric neuroradiology (a-MRI, DWI, MRS, and task-fMRI), pediatric neurologist (with expertise in over 2000 individual age 0-21-year-old’s clinical RS interpretation), and pediatric epileptologist (EEG).

#### EEG

XLTEK System EEG/Sleep Acquisition with Neuroworks and Sleepworks software version 8.5. The EEG/Sleep amplifier utilized is the Brain Monitor Amplifier, with the breakout box “Connex” box. Max sampling rate is 200Hz sampling rate for scalp EEG, and a 256-channel box. A standard array of 19 leads with continuous video EEG (cv-EEG) was interpretated via visual inspection by the pediatric epileptologist. Infants with suspected HIE receive cv-EEG starting on the first day of admission and removed on the day of MRI. The other patients cv-EEG also either occurred on the day of admission.

#### 2.4.2 a-MRI and MRS

All MRI sequences were acquired on a 3T MR scanner (Ingenia, Philips Healthcare, Best, The Netherlands) with 32 channel head coil. T1-weighted turbo field echo whole-brain sequence were obtained with the following parameters: TR (repetition time) 9 msec, TE (echo time) 4 msec, flip angle 8, slice thickness 0.9 mm, and in-plane resolution 0.9 × 0.9 mm. Diffusion weighted (DWI) sequences with corresponding apparent diffusion maps: 2D, 4 mm slice thickness, TR 4.9 msec, TE 0.071 msec, 5 mm space between slices, 124 phase encoding steps, percent phase field of view 96, flip angle 90, and FOV 250×250 mm. H-MRS^1^ data were acquired using a single-voxel point resolved spectroscopy sequence (PRESS; TR□= □ 2□s; TE□= □144□ms; 128 signal averages; voxel size: ∼1 cm^3^) localized to the white matter of the centrum semiovale, and grey matter of the basal ganglia region.Concentrations were estimated using commercially available and fully automated LCModel software (V6.3-1□C). Results yielded the relative concentrations of *N*-acetyl-aspartate (NAA), choline, creatine, lipids, lactate and myoinositol. Interpretation of a-MRI, DWI, and MRS was performed by the pediatric neuroradiologist.

#### 2.4.3 Task-fMRI

Passive movement of the right index finger by the MRI technician was performed with a goal to activate the left sensory motor cortex. SensaVue fMRI equipment (Invivo Corporation, Gainesville, FL, USA) delivered a 5-minute sequence with 8 blocks of activity, 15 seconds of baseline scan discarded, 15s rest, 15s task; TR 3000 msec, TE 35, matrix size 128×128, flip angle 90, slice thickness 4mm, 1.8 × 1.8 × 4 mm voxel, in-plane FOV 230 × 230mm, and ≥ 35 slices, as adjusted to patient head size. The tb-fMRI was visually inspected for expected specific recorded real-time network activation. Resultant SensaVue general linear model difference between activation and baseline images were interpreted by the pediatric neuroradiologist.

#### 2.4.4 RS

T2-weighted images were acquired with a TR 2000 ms, TE 30 ms, matrix size 80 × 80, flip angle 80, number of slices to cover at least supratentorium, slice thickness such that voxel size is 3.4 × 3.4 × 3.4 mm^3^, no slice gap, and inter-leaved top-down acquisition. The number of total volumes was 600, split between two equally timed runs, each approximately 10 minutes. During preprocessing, the first five volumes were deleted to remove T1 saturation effects, high pass filter 100 sec, inter-leaved slice time correction, spatial smoothing 1mm, and motion corrected by MCFLIRT, (Jenkinson et al., 2002) with non-brain structures removed. All subjects had less than 0.5mm motion-induced displacement in any direction. Individual functional scans were registered to the patient’s high-resolution anatomical scan by using linear registration (Jenkinson et al., 2002), with optimization using boundary-based registration (Greve and Fischl, 2009). RS independent component analysis (ICA), a data driven approach, was performed as in prior work (FMRIB MELODIC) (Boerwinkle et al., 2017; Griffanti et al., 2017; Mongerson et al., 2017; Beckmann and Smith, 2004). The results were manually sorted into noise verses neuronal signal. Expert visual interpretation of RS ICA outcomes in patients age 0-21 years with mixed causative pathologies including brain injury have been reported (Boerwinkle et al., 2020; Boerwinkle et al., 2019a; Boerwinkle et al., 2019b; Boerwinkle et al., 2018; Desai et al., 2018; Boerwinkle et al., 2017). Noise components for example, were located outside of the brain, in the cerebral spinal spaces, over major arteries, whereas known RSN were located primarily over the grey matter with well published spatial patterns (Boerwinkle et al., 2017). Normal RSN were those detected with spatial consistency as previously described (e.g. motor, vision, default mode) and with blood oxygenation level dependent signal (BOLD) frequency less than 10 Hz/100 (defined as normal for neonate herein) (Boerwinkle et al., 2019a; Boerwinkle et al., 2017). This was supported by prior work wherein visual inspection of spatial consistency was high compared to co-registered volumetric adult comparison (Linke et al., 2018).

### 2.5 Resting State Networks, Abnormal and Seizure Onset Zone Networks

Detection of RSN in the neonatal population include the major resting state networks, though known maturational changes still occur after this period, especially through the increasing of long-range connectivity. In the neonate’s RS clinical reports, each non-noise RSN is shown with its corresponding BOLD signal time course and associated frequency power spectrum. However, due to need for a data reduction step for the study, with the exception of lack total RSN detection, only the RSN evaluated for study evaluation were: (1) the basal ganglia (BG), due its relationship with frequency of insult in neonates leading to motor disorders; (2) the language/frontal-parietal (Lang-FP), due to the relationship of these networks with cognitive and communication capacities. These were combined to reduce the total number of comparisons, since nuanced differences in language and cognitive outcomes were not evaluated specifically; (3) the default mode (DMN), due to its relationship with consciousness, and outcomes after brain injury. (4) atypical (non-RSN) networks, which were located over grey matter primarily, and did not have features consistent with published RSN or RS noise (Boerwinkle et al., 2019a; Boerwinkle et al., 2017). Atypical networks were further categorized as either consistent with RS-SOZ, low or indeterminate consistency with RS-SOZ, or not consistent with RS-SOZ, according to the spatial and temporal features developed in children with drug resistant epilepsy (DRE) (Boerwinkle et al., 2019a; Boerwinkle et al., 2017). Of note, the timing of actual clinical or electrographic seizures after brain injury is highest in the immediate period (1-3 days), whereas the RS was typically performed by day 5 or after. Also, actively seizing neonates are not transferred for elective MRI. Thus, ongoing clinical seizures did not occur during the scan and seizures electrographic seizures were not measured during or near the time of the scan. Those neonates who did have seizures were continued on anti-seizure medication during the time of scan. Similarly, in children with DRE, findings by RS seemingly consistent with a seizure onset zone are acquired almost entirely during the interictal period and are thus thought to indicate ongoing abnormal network activity that occurs at sub-clinical seizure threshold. Ultimately, these findings have shown to be reliable to direct neurosurgical intervention leading to improved surgical outcomes (Chakraborty et al., 2020; Boerwinkle et al., 2019a; Boerwinkle et al., 2017). However, being that this population does not have the diagnosis of DRE, the meaning of RS SOZ after ABI in those with largely resolved seizures may have more to do with the neuroplastic changes of the injury, rather than correlation with a resolved seizure or prediction of epileptogenicity. Lastly, those subjects with a missing one of these RSN or no RSN detected were noted because the poorest RS connectivity is reported in the few cases after brain death (Kumar et al., 2016; Boly et al., 2009; Han et al., 2006; Jain and DeGeorgia, 2005; Saposnik et al., 2005; Marti-Fabregas et al., 2000; James, 1998; Jorgensen and Malchow-Moller, 1981). Thus, the investigators expected lack of RSN detection to occur more often in those with poorer outcomes, though that would likely be determined only on a descriptive level (though still interesting for informing future prospective trial design) as survival after acute brain injury in neonates is fairly robust.

### 2.6 Statistical Analyses

The distribution of demographics, clinical factors and outcomes were summarized using descriptive statistics, and compared between RSN categories using the Fisher exact (FE) or Kruskal-Wallis test, as appropriate for the data distribution. The magnitude of association was between RSN category scores and factors/outcomes quantified using odds ratio (OR) estimates and corresponding confidence intervals (CI) from ordinal logistic regression analyses (OLR) or multinomial logistic regression models (MLR) when the proportional odds assumption was not valid. Similar analyses examined association of clinical factors and outcomes with findings from EEG, a-MRI, task-fMRI, and MRS modalities. For OLR/MLR models, the predictor ordinal levels were fit as a continuous score. Pairwise agreement between modalities were assessed using the Spearman rank correlation coefficient (CC). All statistical tests were 2-sided, with statistical significance and CI adjusted using a Bonferroni correction to account for multiple test comparisons for each modality (adjusted threshold *P*<.006 for 8 or 9 comparisons per modality; adjusted threshold *P*<.002 for 22 pairwise comparisons among modalities).

## 3. Results

Subjects (N=40) had a mean (standard deviation) gestational age of 37.8 (2.6) weeks, and included 25 (63%) males, 27 (68%) with HIE, and no RS-related safety events. Three (8%) subjects died prior to discharge and one subject was lost to follow-up. Among the 36 (90%) subjects followed, the median age (interquartile range) at outpatient follow-up visit was 31.6 (13.9, 37.6) weeks. Findings of presentation, neurological exam, consciousness, RS, a-MRI, task-fMRI, MRS, EEG, and outcomes during the acute period are shown in **Table 1** and an example of normal versus atypical RS is shown in **Figure 1**.

**Figure 1.**
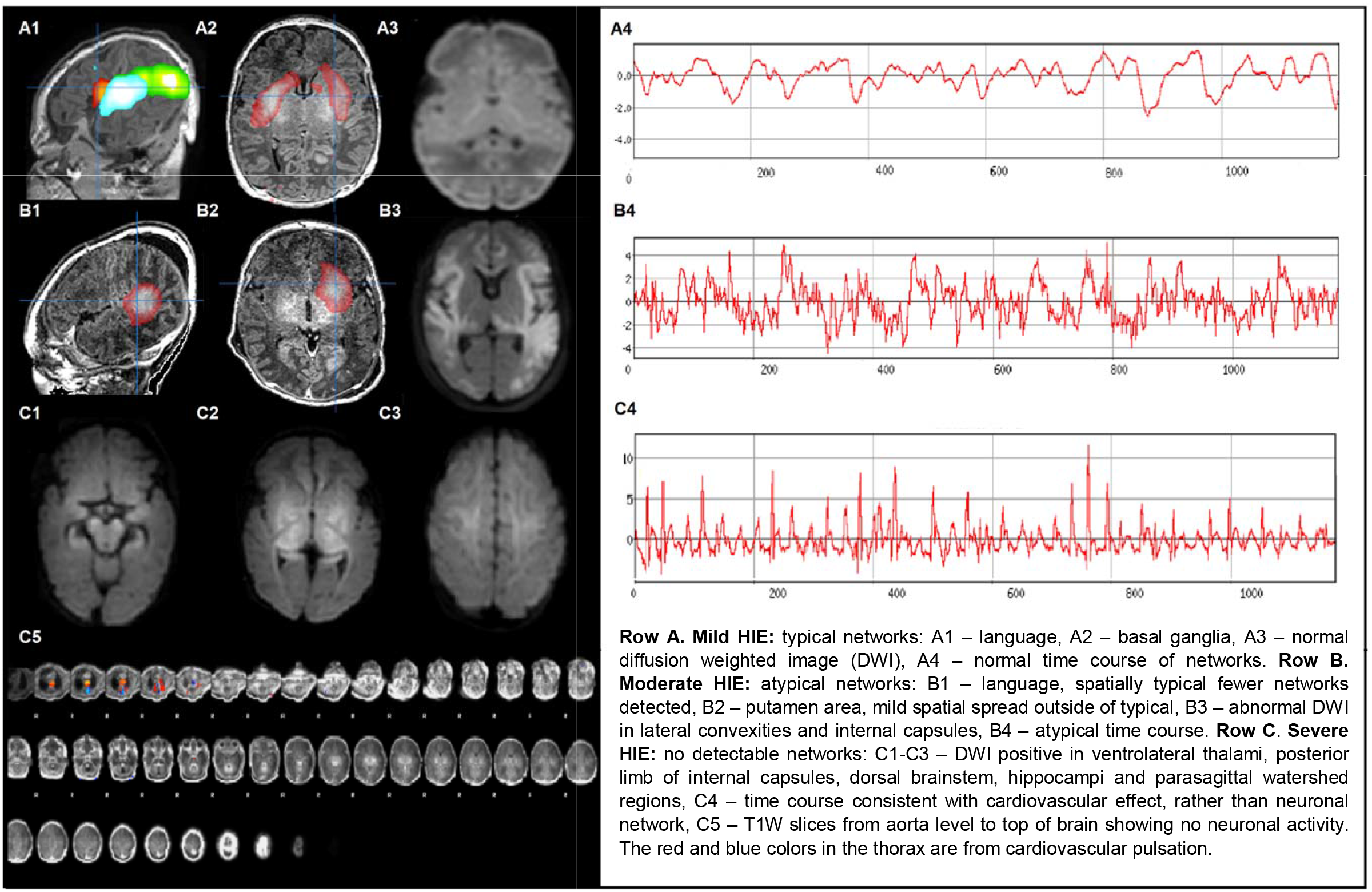
Resting-State Networks (RSN) in Neonates with Mild to Severe Hypoxic Ischemic Encephalopathy (HIE).

All association analyses between tests and clinical factors/outcomes are presented in the supplementary material (**eTables 1-8**), whilst **Tables 2 and 3** presents those associations showing significance after multiple testing correction.

**Table 2:**
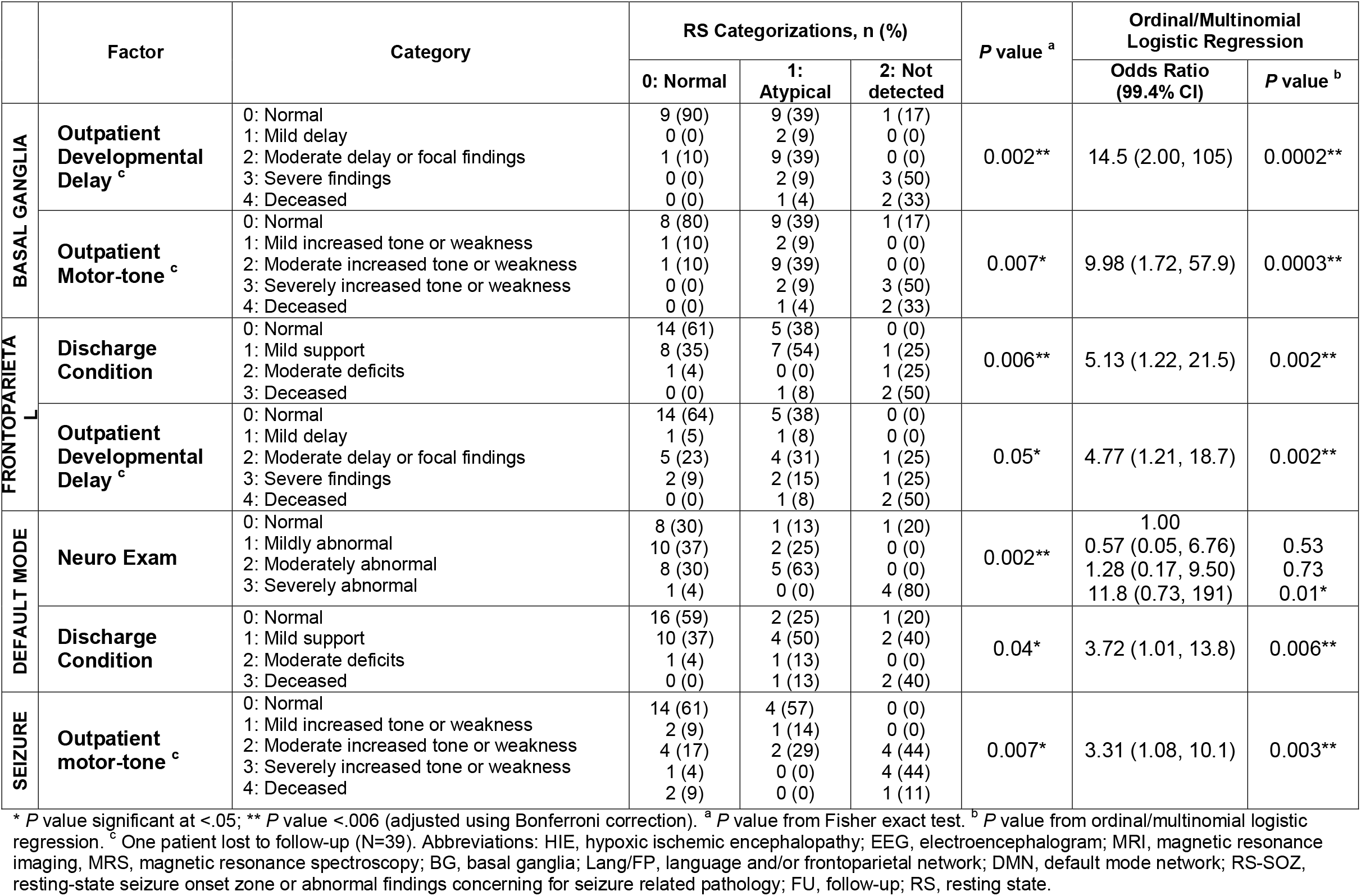
Associations of RSN with Acute Factors and Outcomes in Neonates (N=40)

**Table 3:** Association of Baseline Factors with Acute Tests on Neonates (N=40)

|  | Factor | Category | RS Categorizations, n (%) |  |  |  | P value <sup>a</sup> | Ordinal/Multinomial Logistic Regression |  |
| --- | --- | --- | --- | --- | --- | --- | --- | --- | --- |
|  |  |  | 0: Normal | 1: Mildly abnormal | 2: Moderately abnormal | 3: Severely abnormal |  | Odds Ratio (99.4% CI) | P value <sup>b</sup> |
| Anatomical MRI | HIE (N=27) | 1: Mild<br>2: Moderate<br>3: Severe | 8 (73)<br>2 (18)<br>1 (9) | 5 (63)<br>3 (38)<br>0 (0) | 1 (100)<br>0 (0)<br>0 (0) | 0 (0)<br>2 (29)<br>5 (71) | 0.002** | 3.54 (1.18, 10.7) | 0.002** |
|  | Neuro Exam | 0: Normal<br>1: Mildly abnormal<br>2: Moderately abnormal<br>3: Severely abnormal | 7 (47)<br>6 (40)<br>1 (7)<br>1 (7) | 1 (9)<br>1 (9)<br>9 (82)<br>0 (0) | 1 (20)<br>3 (60)<br>1 (20)<br>0 (0) | 1 (11)<br>2 (22)<br>2 (22)<br>4 (44) | 0.0002** | 2.14 (1.01, 4.56) | 0.006** |
|  | Outpatient Developmental Delay <sup>c</sup> | 0: Normal<br>1: Mild delay<br>2: Moderate delay or focal findings<br>3: Severe findings<br>4: Deceased | 10 (71)<br>1 (7)<br>1 (7)<br>2 (14)<br>0 (0) | 7 (64)<br>0 (0)<br>2 (18)<br>1 (9)<br>1 (9) | 0 (0)<br>1 (20)<br>3 (60)<br>1 (20)<br>0 (0) | 2 (22)<br>0 (0)<br>4 (44)<br>1 (11)<br>2 (22) | 0.02* | 2.15 (0.99, 4.67) | 0.007* |
|  | Seizure | 0: No<br>1: Yes<br>(Deceased/Lost to follow-up, N=4) | 12 (86)<br>2 (14) | 7 (70)<br>3 (30) | 3 (60)<br>2 (40) | 1 (14)<br>6 (86) | 0.02* | 1.0<br>2.94 (1.02, 8.43) | 0.005** |
| EEG |  |  | 0: Normal | 1: Mild background | 2: Seizure | 3: Flat |  |  |  |
|  | Seizure | 0: No<br>1: Yes<br>(Deceased/Lost to follow-up, N=4) | 2 (100)<br>0 (0) | 18 (86)<br>3 (14) | 2 (17)<br>10 (83) | 0 (0)<br>0 (0) | 0.0001** | 1.00<br>30.4 (2.02, 458) | 0.0005** |

<sup>a</sup> P value <.05; <sup>\*\*</sup> P value <.006 (adjusted using Bonferroni correction); <sup>a</sup> P value from Fisher exact test; <sup>b</sup> P value from ordinal/multinomial logistic regression. <sup>c</sup> One patient lost to follow-up (N=39). Abbreviations: HIE, hypoxic ischemic encephalopathy; EEG, electroencephalogram; MRI, magnetic resonance imaging; fMRI, functional MRI; MRS, magnetic resonance spectroscopy.

After multiple testing correction, associations remaining significant for RSN were: BG with outpatient developmental delay (OR, 14.5; 99.4% CI, 2.00-105; *P*<.001) and motor weakness/tone (OR, 9.98; 99.4% CI, 1.72-57.9; *P*<.001); Lang/FP with discharge condition (OR, 5.13; 99.4% CI, 1.22-21.5; *P*=.002) and outpatient developmental delay (OR, 4.77; 99.4% CI, 1.21-18.7; *P=*.002); DMN with discharge condition (OR, 3.72; 99.4% CI, 1.01-13.8; *P=*.006) and neurological exam (*P=*.002(FE); OR, 11.8; 99.4% CI, 0.73-191; *P=*.01(OLR)); and RS-SOZ with motor weakness/tone at follow-up (OR, 3.31; 99.4% CI, 1.08-10.1; *P=*.003) (**Table 2**).

Findings across all test modalities are summarized in supplementary **eTable 9**. Among the non-RS diagnostics (**Table 3**), only a-MRI was associated with neurological exam (OR, 2.14; 99.4% CI, 1.01-4.56; *P*<.001), after multiple testing correction. Outpatient seizure concern was associated with both a-MRI (OR, 2.94; 99.4% CI, 1.02-8.43; *P*=.005) and more strongly with EEG (OR, 30.4; 99.4% CI, 2.02-458; *P*<.001).

Cross-modality correlations significant after multiple testing were detected for a-MRI with EEG (CC, 0.52; 99.8% CI, 0.07-0.80; *P*<.001) and MRS (CC, 0.74; 99.8% CI, 0.22-0.93; *P*<.001) (**Table 4**). While not meeting multiple testing correction criteria, RSN correlation with other diagnostic modalities were suggested only between Lang/FP with task-fMRI and RS-SOZ with a-MRI (**Table 4**).

**Table 4.** Spearman Correlation Coefficient and 99.8% Confidence Intervals (CI) between Resting-State Networks (RSN) and Other Test Modalities

|  | Spearman Correlation Coefficient (99.8% CI)<br>*P value* |  |  |  |  |  |  |
| --- | --- | --- | --- | --- | --- | --- | --- |
| Other Test Modalities | RSN |  |  |  | Acute Test Modalities |  |  |
|  | BG | Lang/FP | DMN | RS-SOZ | EEG | Anatomical MRI | Task-fMRI |
| EEG | 0.22<br>(-0.28, 0.63)<br>$P=0.17$ | 0.20<br>(-0.29, 0.62)<br>$P=0.21$ | 0.21<br>(-0.29, 0.62)<br>$P=0.19$ | 0.20<br>(-0.30, 0.62)<br>$P=0.21$ | - | | - |
| Anatomical MRI | 0.18<br>(-0.32, 0.60)<br>$P=0.27$ | 0.27<br>(-0.34, 0.58)<br>$P=0.08$ | 0.15<br>(-0.34, 0.58)<br>$P=0.34$ | 0.44<br>(-0.04, 0.75)<br>$P=0.004$ | 0.52<br>(0.07, 0.80)<br>$P=0.0004^*$ | - | - |
| Task-fMRI | 0.31<br>(-0.22, 0.69)<br>$P=0.07$ | 0.46<br>(-0.32, 0.63)<br>$P=0.004$ | 0.20<br>(-0.32, 0.63)<br>$P=0.23$ | -0.12<br>(-0.58, 0.39)<br>$P=0.47$ | 0.25<br>(-0.28, 0.67)<br>$P=0.14$ | 0.38<br>(-0.14, 0.73)<br>$P=0.02$ | - |
| MRS | 0.02<br>(-0.61, 0.64)<br>$P=0.93$ | 0.33<br>(-0.49, 0.72)<br>$P=0.14$ | 0.19<br>(-0.49, 0.72)<br>$P=0.41$ | 0.18<br>(-0.50, 0.72)<br>$P=0.43$ | 0.37<br>(-0.32, 0.81)<br>$P=0.09$ | 0.74<br>(0.22, 0.93)<br>$P=<0.0001^*$ | 0.57<br>(-0.12, 0.89)<br>$P=0.008$ |

\* $P$ at $<.002$ meeting multiple testing criteria. Abbreviations: EEG, electroencephalogram; MRI, magnetic resonance imaging; fMRI, functional MRI; MRS, magnetic resonance spectroscopy; BG, basal ganglia network; Lang/FP, language and/or frontoparietal network; DMN, default mode network; RS-SOZ, resting-state seizure onset zone or abnormal findings concerning for seizure related pathology; RSN, resting-state networks.

Among the three patients who died, BG, Lang/FP, and DMN were atypical in one and not detected in two patients (**Figure 1**). The three that died while in the hospital had insults due to traumatic brain injury (TBI), severe HIE, and placental abruption. The first two neonates (TBI and severe HIE) had no detectable RSNs. The first’s exam progressed to findings consistent with brain death (BD), (**Figure 1**) although apnea and cold caloric tests not performed. In all three, the medical condition was determined to be terminal or futile after RS, and then families and team elected WLST. Associations with mortality were stronger for RS (though not significant after correction for multiple testing) than for a-MRI, task-fMRI, MRS, and EEG (**eTables 1-8**). Associations with consciousness were suggested and comparable for Lang/FP, MRS, DMN, a-MRI (**eTables 1-8**).

## 4. Discussion

RSN detection indicated good signal quality was obtained safety in this tertiary neonatal intensive care unit ABI severity environment, thus, worthy of consideration for care. In this study, RS was also consistent with prior findings of emerging before birth (Doria et al., 2010) and increasing rapidly during this early developmental period (Gao et al., 2015; Gao et al., 2011).

In this cohort of neonates with common ABI etiologies, IBFN via RS was associated with neurological exam and outcomes, suggesting the connectivity degradation was physiological and related to the injury, rather than causes such as theorized early developmental RSN poor signal, ICU-related therapies, or artifact. These RS findings, pointing to ABI impacting the neural network function and plasticity, are consistent with results from other modalities evaluating adults and children with ABI (Smyser and Neil, 2015).

### 4.1 Incremental Outcomes

The *incremental* degradation of RSN connectivity was associated with worsened *outcomes, and findings specific to different networks*. The most severe RSN finding, lack of either the Lang-FP or DMN, made it more likely that upon discharge, there would be severely abnormal exam findings or death. Similarly, worse BG and RS-SOZ were related to marked outpatient developmental delay or motor delay, respectively, or death. Incrementally worse a-MRI was associated with *worsening* acute factors, and *overall* outpatient development at follow-up. This difference may be due to the lack of blinding with cross-informed sub-specialist interpretations in the acute period being greater with more substantiated tests. However, the association with outcomes from these prospectively documented clinical notes may be informative, as suggested by Linke et al (Linke et al., 2018).

### 4.2 Developmental Outcomes

The BG and RS-SOZ were associated with motor outcome, suggesting specific insight into early motor skills and correspondence to subcortical-dominated HIE neonatal pathology (Douglas-Escobar and Weiss, 2015). Correspondingly, the spatially broader and cortical RSN, Lang-FP and DMN, had association with more generalized developmental outcomes. Supportively, FP is the earliest to stabilize in connectivity and has predictive capacity in individually unique cognitive capacities (Wang et al., 2021; Doria et al., 2010). This may help explain the cognitive and other cortically localized outcomes known to occur after HIE, though longer follow up is indicated (Pouppirt et al., 2021; Douglas-Escobar and Weiss, 2015; van Kooij et al., 2010).

### 4.3 Acute EEG and Seizure Outcomes

Only the EEG and a-MRI were specifically related to seizure outcomes, whereas the RS-SOZ and seizure outcome association was weaker. And, the RS-SOZ was *not* associated with acute EEG. The reasons for this could be multifactorial. (1) The study design in determination of seizure outcome was weak, as it was based largely on parent description of repeat events which can be subtle and difficult to discern in neonates or practioners style of continued antiseizure at the time of follow up visit. (2) The seizure captured on EEG and often clinically resolved by the time of RS may reflect a true phenomenon of disparately timed tests capturing the actual evolution of the brain condition in ABI. Supportively, most ABI seizures occur 0-72 hours after insult, (Lynch et al., 2015) whereas RS capture was after 72 hours. (3) Conversely negative EEG could possibly still occur despite deep-source seizures, whereas RS-SOZ has no such spatial limitation. This may explain why the RS-SOZ unadjusted association was highest with a-MRI and subsequent developmental delay and motor assessment (supplementary material (**eTables 4**) (Lopes et al., 2012; Wurina et al., 2012).

### 4.4 Acute Exam, Consciousness, and Mortality

We demonstrate consistency across exams and tests, which may affect WLST, and suggests potential for impact on acute period care. As such, the minimal IBNF for survival and meaningful brain recovery is a key factor. Unadjusted associations suggest that the minimum IBFN by RS for *survival* may be above complete lack of RSN detection, or lack of Lang-FP detection (75% mortality), and subtler findings may change due to neuroplasticity (Boerwinkle et al., 2019a; Boerwinkle, 2018). Also, re-emergence of RSN is unknown in ABI, though occurs in the anesthetized. This is unknown in neonates, and therefore, is a limitation of care with ethical implications. The Lang-FP connection to mortality could be reflective of this network combination covering a very broad spatial territory of parenchyma. This may mean a large portion of the brain is in the irreversible cell-death cascade, which can trigger a more systemic destabilization, and death. Otherwise, RSN and a-MRI associated outcomes included death, suggesting the linkage to systemic health and death. Thus, serial longitudinal RS after ABI is indicated for evaluation of re-emergence and increased minimal IBNF definition.

The DMN was the only network with statistically significant *overall* association with the acute neurologic exam, which included evaluation of consciousness, as best it could be determined by responsiveness and eye opening to environmental stimuli. This suggests that markers of consciousness in neonates with ABI across the full severity range may be related to ongoing supratentorial default mode network activity, rather than a mere brainstem mediated phenomenon. This network outcome association are supported by prior work: Lang-FP (Guo et al., 2016; Boly et al., 2009) and DMN (Threlkeld et al., 2018; Kondziella et al., 2017) in other ABI populations, which were associated with acute neurological findings. Comparatively, among the non-RSN tests, a-MRI was also associated with the neurological exam.

*HIE severity* was associated with a-MRI, but not RSNs. This could be due to the study design wherein the HIE severity was ultimately determined after all diagnostics and acute exams, at discharge, thus is not blinded, and may be largely based on the a-MRI findings. A blinded prospective study would solve the question.

### 4.5 Cross-modality Observations

The cross-modality severity consistency has supportive and unique results in this real-world all-ABI-comer neonatal population. Namely, the BG, a-MRI, and MRS rates of being atypical were commensurately. Contrastingly, the cortical RSNs had similar but lower atypical rates. Thus, subcortical, more so than cortical, network abnormality is as expected in an HIE-rich cohort (de Vries and Groenendaal, 2010). Notably, non-RS tests had higher abnormality rates, yet the RSN had higher frequency of association with outcomes, suggesting unique information from RSNs with development.

Inter-modality disparities are expected and reflect different physiological signals. Thus, the higher rate of cortical RSN normality is in-line with task-fMRI, measuring cortical motor function. Similarly, MRS, measuring deep-brain location metabolism, was more often abnormal in this HIE-rich population. Hence the disparities are consistent and triangulate on different and localizing aspects of the ABI etiology.

### 4.6 Limitations and Further Study

A prospective approach with standardized exams and outcomes measures are needed. Similarly, validated neonatal consciousness and other acute factor and outcome measures are necessary for mixed pathology/full range severity populations.

RSN requires interpretation, prompting the blinded study design. Further means of minimizing bias by developing automated RSN categorization may help (Chakraborty et al., 2020; Griffanti et al., 2017; Griffanti et al., 2014).

In neonates with ABI, network pathology localization is not well-characterized and data-driven approaches, such as ICA may best be suited for localization of network pathology. ICA has level 1 evidence for diagnostic testing (OCEBM Levels of Evidence Working Group) in children and adults for characterizing normal and pathological networks in drug resistant epilepsy and improving outcomes (Chakraborty et al., 2020), however, method comparison may be of value.

## Conclusions

This study provides level 3 evidence (OCEBM Levels of Evidence Working Group) that RSN are associated with outcomes in neonates with ABI. Specific networks’ degradation was linked to different worsened outcomes. The basal ganglia and seizure onset zone networks were associated with motor outcomes. The broad language/cognitive region networks were associated with developmental delay. Discharge condition, which includes mortality, was associated with default mode and language/cognitive networks. Lack of all studied networks only occurred in those who did not survive.

## Data Availability

All data produced in the present work are contained in the manuscript or supplementary material.

## Abbreviations

ABI: acute brain injury
IBNF: integrated brain network function
DoC: disorders of consciousness
WLST: withdrawal of life sustaining therapy determinations
EEG: electroencephalogram
fMRI: functional magnetic resonance imaging
RSN: resting state network(s)
DMN: default mode network
FP: frontal-parietal network
Lang-FP: language/frontal-parietal network
HIE: hypoxic ischemic encephalopathy
a-MRI: anatomical MRI
BD: brain death
RS: resting state functional magnetic resonance imaging
MRS: magnetic resonance spectroscopy
cv-EEG: continuous video EEG
TR: repetition time
TE: echo time
ICA: independent component analysis
BOLD: blood oxygenation level dependent signal
DRE: drug resistant epilepsy
FE: Fisher exact
OR: odds ratio
CI: confidence intervals
OLR: ordinal logistic regression analyses
MLR: multinomial logistic regression models
CC: correlation coefficient
TBI: traumatic brain injury

## Acknowledgements

The authors are grateful for the patients included in this study. We would like to give a special thank you to their families and our colleagues involved in their care at Phoenix Children’s Hospital. Institutional Review Board (IRB # 20-331) at Phoenix Children’s Hospital approved this study. This research did not receive any specific grant from funding agencies in the public, commercial, or not-for-profit sectors.

## Notes

### Competing Interest Statement

The authors have declared no competing interest.

### Funding Statement

This study did not receive any funding

### Author Declarations

Ethics committee/IRB of Phoenix Childrens Hospital gave eithial approval for this work.

