## Supplementary Material for "Association of Network Connectivity via Resting State Functional MRI with Consciousness, Mortality, and Outcomes in Neonatal Acute Brain Injury"

**Supplemental Material**

**Modality Details**

Acute testing included continuous electroencephalogram (EEG), anatomic magnetic resonance imaging (a-MRI), task-based functional MRI (Task-fMRI), resting state (RS) MRI, and magnetic resonance spectroscopy (MRS).

EEG was performed throughout the acute period. EEGs were recorded using Natus (Middleton, WI U.S.A.) Xltec equipment, with a sampling rate of 200 Hz, using a standard neonatal montage^1^. A board-certified pediatric neurophysiologist identified events as seizures based on standard EEG interpretation. Seizures on traditional EEG were defined as rhythmic discharges evolving in frequency, amplitude, morphology, and/or spatial distribution, lasting at least 10 seconds.

MRI were acquired on a 3 Tesla MRI (Ingenuity, Philips Medical Systems, Best, Netherlands) equipped with a 32-channel head coil. RS parameters included TR 2000 ms, TE 30 ms, matrix size 80 x 80, flip angle 80, number of slices 46, slice thickness 3.4 mm with no gap, in plane resolution 3 x 3 mm, and number of total volumes 600 lasting 20 minutes (two, 10-minute runs), as prior^2^. A T2 weighted-3D whole-brain sequence, designed for neonates, was obtained for anatomical reference with TR 2.5 ms, TE 331 ms, flip angle 90, slice thickness 0.9 mm, and in plane resolution of 0.8 x 0.8 mm.

Subjects who remained still also received a 5-minute passive task-MRI, with same sequence parameters as the RS (Invivo SensaVue system; Gainesville, FL, USA). A visual paradigm of a kaleidoscope pattern projection was alternated with fixed cross visual presentation. A positive test was statistically significant activation within the occipital cortices. Passive motor finger tapping stimulation by a technician moving the patient’s index finger was also performed. A positive test result was statistically significant motor cortex activation. Images were processed using DynaSuite Neuro Software (Invivo Corporation, Gainesville, FL), which involves motion correction, co-registration of the fMRI data set with T1 weighted magnetic resonance images and generalized linear model processing of block paradigm type designs.

Standard RS preprocessing steps were applied: non-brain structures were removed, the first 5 volumes were deleted to remove T1 saturation effects, high pass filter 100 s, inter-leaved slice time correction, minimal spatial smoothing at 1 mm, and motion corrected by MCFLIRT^3, 4^. All subjects had less than 0.5 mm motion-induced displacement in any direction. Individual functional scans were registered to the patient’s high-resolution anatomical scan using linear registration^5^, with optimization using boundary-based registration^6^. Patient registration to a standard template was not conducted, nor was spatial smoothing nor mean signal regression.

Independent component analysis (ICA) is a mathematical process that analyzes the raw RS signal and separates it into oscillating sub-signals. These sub-signals (called independent components – IC) can be either from brain networks or from noise - such as from the MRI scanner, respiration, cardiovascular activity, and cerebrospinal fluid pulsation. Determining what is noise from the brain networks can be challenging and is described in the literature^4^. Each brain network oscillates in its oxygen concentration separately from the other brain networks, which is what makes each brain network separately detectable from the others. Thus, the normal time course of this oscillation pattern in functional gray matter is typically smooth and slow, and usually less than 2 Hz. In comparison, brain networks that are abnormal often have fast and erratic blood oxygen dependent (BOLD) time courses^4^. The total number of detected ICs was determined using the established automated dimensionality estimate that utilizes a Bayesian approach^7^. As such, the threshold of the ICA maps was set by the standard local false discovery rate for IC detection at p<.05 ^7^.

Expertise level in pediatric RS interpretation is available at our institution, with over 1500 individual patient RSs interpreted, with age newborn to 21 years. Automated RS interpretation has not been clinically validated in neonates. Like interpretation of a-MRIs, RS interpretation is performed by those with clinical experience and is, therefore, limited by the same levels and types of subjective bias^8, 9^. The approach, described by Boerwinkle, has the largest reported level of reported clinical application in children^4, 10^. This was previously applied to an intractable pediatric epilepsy population, largely secondary to prior brain injuries and congenital etiologies.

**eTable 1: Association of Resting State Basal Ganglia (RS-BG) with Outcomes.**

| **Factor** | **RS-BG** | | | **P**  **value ^a^** | **Ordinal/Multinomial**  **Logistic Regression** | |
| --- | --- | --- | --- | --- | --- | --- |
|  | **0: Normal**  **(N=10)** | **1: Atypical**  **(N=24)** | **2: Not detected**  **(N=6)** |  | **Odds Ratio**  **(99.4% CI)** | **P**  **value ^b^** |
| **HIE**, N (%)  0: No  1: Yes | 4 (40)  6 (60) | 8 (33)  16 (67) | 1 (17)  5 (83) | 0.72 | 1.00  1.66 (0.36, 7.70) | 0.36 |
| **HIE**, N (%)  1: Mild  2: Moderate  3: Severe | 6 (100)  0 (0)  0 (0) | 6 (38)  6 (38)  4 (25) | 2 (40)  1 (20)  2 (40) | 0.09 | 4.59 (0.68, 31) | 0.03* |
| **Neuro Exam**, N (%)  0: Normal  1: Mildly abnormal  2: Moderately abnormal  3: Severely abnormal | 2 (20)  4 (40)  4 (40)  0 (0) | 7 (29)  8 (33)  8 (33)  1 (4) | 1 (17)  0 (0)  1 (17)  4 (67) | 0.02* | 1.00  0.44 (0.04, 4.30)  0.62 (0.07, 5.77)  32.1 (0.73, >99) | 0.32  0.56  0.01* |
| **Consciousness (Day 0-5)**, N (%)  0: Normal  1: Irritable but arouses easily  2: Wakes up only to painful stimulation  3: Coma  4: Episodically arousable | 2 (20)  5 (50)  2 (20)  1 (10)  0 (0) | 6 (25)  10 (42)  7 (29)  0 (0)  1 (4) | 1 (17)  1 (17)  2 (33)  2 (33)  0 (0) | 0.32 | 1.67 (0.46, 6.07) | 0.27 |
| **Death**, N (%)  0: No  1: Yes | 10 (100)  0 (0) | 23 (96)  1 (4) | 4 (67)  2 (33) | 0.10 | 1.00  12.62 (0.38, 414) | 0.05* |
| **Discharge Condition**, N (%)  0: Normal  1: Mild support  2: Moderate deficits  3: Deceased | 5 (50)  5 (50)  0 (0)  0 (0) | 13 (54)  10 (42)  0 (0)  1 (4) | 1 (17)  1 (17)  2 (33)  2 (33) | 0.02* | 1.00  0.87 (0.15, 4.95)  >99 (<0.01, >99)  22.9 (0.47, >99) | 0.83  0.99  0.03* |
| **Outpatient Development**, N (%)  0: Normal  1: Mild delay  2: Moderate delay or focal finding on exam  3: Severe findings  4: Deceased | 9 (90)  0 (0)  1 (10)  0 (0)  0 (0) | 9 (39)  2 (9)  9 (39)  2 (9)  1 (4) | 1 (17)  0 (0)  0 (0)  3 (50)  2 (33) | 0.002** | 14.5 (2.0, 105) | 0.0002** |
| **Outpatient Motor-Tone**, N (%)  0: Normal  1: Mild increased tone or weakness  2: Moderate increased tone or weakness  3: Severely increased tone or weakness  4: Deceased | 8 (80)  1 (10)  1 (10)  0 (0)  0 (0) | 9 (39)  2 (9)  9 (39)  2 (9)  1 (4) | 1 (17)  0 (0)  0 (0)  3 (50)  2 (33) | 0.007 | 9.98 (1.72, 57.9) | 0.0003** |

* P value significant at <.05 (not adjusted using Bonferroni correction). ** P value significant or marginally significant at <.006 (adjusted using Bonferroni correction). ^a^ P value from Fisher exact test. ^b^ P value from ordinal/multinomial logistic regression.

**eTable 2: Association of Resting State (RS) Language and/or Frontal-parietal (FP) with Outcomes**

| **Factor** | **RS-Language and/or FP** | | | **P value ^a^** | **Ordinal/Multinomial**  **Logistic Regression** | |
| --- | --- | --- | --- | --- | --- | --- |
|  | 0: Normal  (N=23) | 1: Atypical  (N=13) | 2: Not detected  (N=4) |  | **Odds Ratio**  **(99.4% CI)** | **P value ^b^** |
| **HIE**, N (%)  0: No  1: Yes | 7 (30)  16 (70) | 4 (31)  9 (69) | 2 (50)  2 (50) | 0.78 | 1.00  0.75 (0.19, 2.91) | 0.56 |
| **HIE**, N (%)  1: Mild  2: Moderate  3: Severe | 11 (69)  2 (13)  3 (19) | 3 (33)  4 (44)  2 (22) | 0 (0)  1 (50)  1 (50) | 0.12 | 3.13 (0.58, 17.0) | 0.06 |
| **Neuro Exam**, N (%)  0: Normal  1: Mildly abnormal  2: Moderately abnormal  3: Severely abnormal | 7 (30)  8 (35)  7 (30)  1 (4) | 3 (23)  3 (23)  5 (38)  2 (15) | 0 (0)  1 (25)  1 (25)  2 (50) | 0.35 | 2.72 (0.77, 9.56) | 0.03* |
| **Consciousness (Day 0-5)**, N (%)  0: Normal  1: Irritable but arouses easily  2: Wakes up only to painful stimulation  3: Coma  4: Episodically arousable | 7 (30)  9 (39)  6 (26)  1 (4)  0 (0) | 2 (15)  6 (46)  4 (30)  0 (0)  1 (8) | 0 (0)  1 (25)  1 (25)  2 (50)  0 (0) | 0.19 | 2.67 (0.76, 9.39) | 0.03* |
| **Death**, N (%)  0: No  1: Yes | 23 (100)  0 (0) | 12 (92)  1 (8) | 2 (50)  2 (50) | 0.008 | 1.00  16.4 (0.54, 498) | 0.02* |
| **Discharge Condition**, N (%)  0: Normal  1: Mild support  2: Moderate deficits  3: Deceased | 14 (61)  8 (35)  1 (4)  0 (0) | 5 (38)  7 (54)  0 (0)  1 (8) | 0 (0)  1 (25)  1 (25)  2 (50) | 0.006** | 5.13 (1.22, 21.51) | 0.002** |
| **Outpatient Development**, N (%)  0: Normal  1: Mild delay  2: Moderate delay or focal finding on exam  3: Severe findings  4: Deceased | 14 (64)  1 (5)  5 (23)  2 (9)  0 (0) | 5 (38)  1 (8)  4 (31)  2 (15)  1 (8) | 0 (0)  0 (0)  1 (25)  1 (25)  2 (50) | 0.05* | 4.77 (1.21, 18.7) | 0.002** |
| **Outpatient Motor-Tone**, N (%)  0: Normal  1: Mild increased tone or weakness  2: Moderate increased tone or weakness  3: Severely increased tone or weakness  4: Deceased | 12 (55)  2 (9)  6 (27)  2 (9)  0 (0) | 5 (38)  1 (8)  4 (31)  2 (15)  1 (8) | 1 (25)  0 (0)  0 (0)  1 (25)  2 (50) | 0.16 | 3.35 (0.92, 12.1) | 0.01* |

* P value significant at <.05 (not adjusted using Bonferroni correction). ** P value significant or marginally significant at <.006 (adjusted using Bonferroni correction). ^a^ P value from Fisher exact test; ^b^ P value from ordinal/multinomial logistic regression.

**eTable 3: Association of Resting State (RS) Default Mode Network with Outcomes**

| **Factor** | **RS-Default Mode Network** | | | **P value ^a^** | **Ordinal/Multinomial**  **Logistic Regression** | |
| --- | --- | --- | --- | --- | --- | --- |
|  | **0: Normal**  **(N=27)** | **1: Atypical**  **(N=8)** | **2: Not detected**  **(N=5)** |  | **Odds Ratio**  **(99.4% CI)** | **P value ^b^** |
| **HIE**, N (%)  0: No  1: Yes | 9 (33)  18 (67) | 2 (25)  6 (75) | 2 (40)  3 (60) | 1.00 | 1.00  0.97 (0.26, 3.58) | 0.94 |
| **HIE**, N (%)  1: Mild  2: Moderate  3: Severe | 12 (67)  3 (17)  3 (17) | 1 (17)  4 (67)  1 (17) | 1 (33)  0 (0)  2 (67) | 0.03* | 3.32 (0.68, 16.3) | 0.04* |
| **Neuro Exam**, N (%)  0: Normal  1: Mildly abnormal  2: Moderately abnormal  3: Severely abnormal | 8 (30)  10 (37)  8 (30)  1 (4) | 1 (13)  2 (25)  5 (63)  0 (0) | 1 (20)  0 (0)  0 (0)  4 (80) | 0.002** | 1.00  0.57 (0.05, 6.76)  1.28 (0.17, 9.50)  11.8 (0.73, 191) | 0.53  0.73  0.01* |
| **Consciousness (Day 0-5)**, N (%)  0: Normal  1: Irritable but arouses easily  2: Wakes up only to painful stimulation  3: Coma  4: Episodically arousable | 7 (26)  12 (44)  7 (26)  1 (4)  0 (0) | 2 (25)  2 (25)  3 (38)  0 (0)  1 (12) | 0 (0)  2 (40)  1 (20)  2 (40)  0 (0) | 0.16 | 2.29 (0.71, 7.45) | 0.05* |
| **Death**, N (%)  0: No  1: Yes | 27 (100)  0 (0) | 7 (88)  1 (13) | 3 (60)  2 (40) | 0.01 | 1.00  10.1 (0.56, 180) | 0.03* |
| **Discharge Condition**, N (%)  0: Normal  1: Mild support  2: Moderate deficits  3: Deceased | 16 (59)  10 (37)  1 (4)  0 (0) | 2 (25)  4 (50)  1 (13)  1 (13) | 1 (20)  2 (40)  0 (0)  2 (40) | 0.04* | 3.72 (1.01, 13.8) | 0.006** |
| **Outpatient Development**, N (%)  0: Normal  1: Mild delay  2: Moderate delay or focal finding on exam  3: Severe findings  4: Deceased | 15 (58)  1 (4)  8 (31)  2 (8)  0 (0) | 3 (38)  0 (0)  2 (25)  2 (25)  1 (13) | 1 (20)  1 (20)  0 (0)  1 (20)  2 (40) | 0.02* | 1.00  4.43 (0.28, 70.6)  0.77 (0.08, 7.30)  3.25 (0.42, 24.9)  15.6 (0.61, 396) | 0.14  0.75  0.11  0.02* |
| **Outpatient Motor-Tone**, N (%)  0: Normal  1: Mild increased tone or weakness  2: Moderate increased tone or weakness  3: Severely increased tone or weakness  4: Deceased | 15 (58)  1 (4)  8 (31)  2 (8)  0 (0) | 2 (25)  1 (13)  2 (25)  2 (25)  1 (13) | 1 (20)  1 (20)  0 (0)  1 (20)  2 (40) | 0.02* | 1.00  5.38 (0.43, 67.2)  0.90 (0.08, 9.41)  3.92 (0.45, 34.2)  19.4 (0.66, 567) | 0.07  0.90  0.08  0.02* |

* P value significant at <.05 (not adjusted using Bonferroni correction). ** P value significant or marginally significant at <.006 (adjusted using Bonferroni correction). ^a^ P value from Fisher exact test. ^b^ P value from ordinal/multinomial logistic regression.

**eTable 4: Association of Resting State (RS) Seizure Onset Zone or Abnormal Findings Concerning for Seizure with Outcomes**

| **Factor** | **RS-Seizure Onset Zone or Abnormal Findings Concerning for Seizure** | | | **P value ^a^** | **Ordinal/Multinomial**  **Logistic Regression** | |
| --- | --- | --- | --- | --- | --- | --- |
|  | **0: Normal**  **(N=24)** | **1: Some Concern for Seizure (N=7)** | **2: High Concern for Seizure**  **(N=9)** |  | **Odds Ratio**  **(99.4 CI)** | **P value ^b^** |
| **HIE**, N (%)  0: No  1: Yes | 8 (33)  16 (67) | 2 (29)  5 (71) | 3 (33)  6 (67) | 1.00 | 1.00  1.02 (0.32, 3.22) | 0.96 |
| **HIE**, N (%)  1: Mild  2: Moderate  3: Severe | 11 (69)  3 (19)  2 (13) | 1 (20)  1 (20)  3 (60) | 2 (33)  3 (50)  1 (17) | 0.09 | 1.92 (0.54, 6.81) | 0.15 |
| **Neuro Exam**, N (%)  0: Normal  1: Mildly abnormal  2: Moderately abnormal  3: Severely abnormal | 7 (30)  9 (38)  6 (25)  2 (8) | 1 (14)  2 (29)  3 (43)  1 (14) | 2 (22)  1 (11)  4 (44)  2 (22) | 0.64 | 1.79 (0.66, 4.90) | 0.10 |
| **Consciousness (Day 0-5)**, N (%)  0: Normal  1: Irritable but arouses easily  2: Wakes up only to painful stimulation  3: Coma  4: Episodically arousable | 5 (21)  12 (50)  5 (21)  2 (8)  0 (0) | 2 (29)  1 (14)  3 (43)  1 (14)  0 (0) | 2 (22)  3 (33)  3 (33)  0 (0)  1 (11) | 0.46 | 1.00  0.69 (0.16, 3.04)  1.23 (0.28, 5.42)  0.55 (0.04, 8.71)  >99 (<0.01, >99) | 0.48  0.70  0.54  0.99 |
| **Death**, N (%)  0: No  1: Yes | 22 (92)  2 (8) | 7 (100)  0 (0) | 8 (89)  1 (11) | 1.00 | 1.00  1.07 (0.14, 7.90) | 0.93 |
| **Discharge Condition**, N (%)  0: Normal  1: Mild support  2: Moderate deficits  3: Deceased | 12 (50)  9 (3)  1 (4)  2 (9) | 4 (57)  3 (43)  0 (0)  0 (0) | 3 (33)  4 (44)  1 (11)  1 (11) | 0.95 | 1.32 (0.47, 3.65) | 0.45 |
| **Outpatient Development**, N (%)  0: Normal  1: Mild delay  2: Moderate delay or focal finding on exam  3: Severe findings  4: Deceased | 14 (61)  1 (4)  5 (22)  1 (4)  2 (9) | 5 (71)  0 (0)  2 (29)  0 (0)  0 (0) | 0 (0)  1 (11)  3 (33)  4 (44)  1 (11) | 0.007* | 2.74 (0.93, 8.06) | 0.009* |
| **Outpatient Motor-Tone**, N (%)  0: Normal  1: Mild increased tone or weakness  2: Moderate increased tone or weakness  3: Severely increased tone or weakness  4: Deceased | 14 (61)  2 (9)  4 (17)  1 (4)  2 (9) | 4 (57)  1 (14)  2 (29)  0 (0)  0 (0) | 0 (0)  0 (0)  4 (44)  4 (44)  1 (11) | 0.007* | 3.31 (1.08, 10.1) | 0.003** |
| **Concern for Seizure**, N (%)  0: No  1: Yes  (Deceased/lost to follow-up, N=4) | 17 (81)  4 (19) | 3 (43)  4 (57) | 3 (38)  5 (63) | 0.04* | 2.83 (0.78, 10.22) | 0.02* |

* P value significant at <.05 (not adjusted using Bonferroni correction). ** P value significant or marginally significant at <.006 (adjusted using Bonferroni correction). ^a^ P value from Fisher exact test; ^b^ P value from ordinal/multinomial logistic regression.

**eTable 5: Association of Baseline Factors with Anatomical MRI.**

| **Factor** | **Anatomical MRI** | | | | **P value ^a^** | **Ordinal/Multinomial Logistic Regression** | |
| --- | --- | --- | --- | --- | --- | --- | --- |
|  | **0: Normal (N=15)** | **1: Mildly abnormal (N=11)** | **2: Moderately abnormal (N=5)** | **3: Severely abnormal (N=9)** |  | **Odds Ratio**  **(99.4% CI)** | **P**  **value ^b^** |
| **HIE**, N (%)  0: No  1: Yes | 4 (27)  11 (73) | 3 (27)  8 (73) | 4 (80)  1 (20) | 2 (22)  7 (78) | 0.14 | 1.00  0.89 (0.40, 1.96) | 0.69 |
| **HIE**, N (%)  1: Mild  2: Moderate  3: Severe | 8 (73)  2 (18)  1 (9) | 5 (63)  3 (38)  0 (0) | 1 (100)  0 (0)  0 (0) | 0 (0)  2 (29)  5 (71) | 0.002** | 3.54 (1.18, 10.68) | 0.002** |
| **Neuro Exam**, N (%)  0: Normal  1: Mildly abnormal  2: Moderately abnormal  3: Severely abnormal | 7 (47)  6 (40)  1 (7)  1 (7) | 1 (9)  1 (9)  9 (82)  0 (0) | 1 (20)  3 (60)  1 (20)  0 (0) | 1 (11)  2 (22)  2 (22)  4 (44) | 0.0002** | 2.14 (1.01, 4.56) | 0.006** |
| **Consciousness (Day 0-5)**, N (%)  0: Normal  1: Irritable but arouses easily  2: Wakes up only to painful stimulation  3: Coma  4: Episodically arousable | 5 (33)  8 (53)  2 (13)  0 (0)  0 (0) | 1 (9)  4 (36)  4 (36)  1 (9)  1 (9) | 2 (40)  2 (40)  1 (20)  0 (0)  0 (0) | 1 (11)  2 (22)  4 (44)  2 (22)  0 (0) | 0.30 | 1.70 (0.83, 3.48) | 0.04* |
| **Death**, N (%)  0: No  1: Yes | 15 (100)  0 (0) | 10 (91)  1 (9) | 5 (100)  0 (0) | 7 (78)  2 (22) | 0.14 | 1.00  2.57 (0.47, 13.9) | 0.12 |
| **Discharge Condition**, N (%)  0: Normal  1: Mild support  2: Moderate deficits  3: Deceased | 8 (53)  6 (40)  1 (7)  0 (0) | 6 (55)  4 (36)  0 (0)  1 (9) | 2 (40)  3 (60)  0 (0)  0 (0) | 3 (3)  3 (3)  1 (11)  2 (22) | 0.72 | 1.48 (0.72, 3.06) | 0.14 |
| **Outpatient Development**, N (%)  0: Normal  1: Mild delay  2: Moderate delay or focal finding on exam  3: Severe findings  4: Deceased | 10 (71)  1 (7)  1 (7)  2 (14)  0 (0) | 7 (64)  0 (0)  2 (18)  1 (9)  1 (9) | 0 (0)  1 (20)  3 (60)  1 (20)  0 (0) | 2 (22)  0 (0)  4 (44)  1 (11)  2 (22) | 0.02* | 2.15 (0.99, 4.67) | 0.007** |
| **Outpatient Motor-Tone**, N (%)  0: Normal  1: Mild increased tone or weakness  2: Moderate increased tone or weakness  3: Severely increased tone or weakness  4: Deceased | 8 (57)  3 (21)  1 (7)  2 (14)  0 (0) | 7 (64)  0 (0)  2 (18)  1 (9)  1 (9) | 1 (20)  0 (0)  3 (60)  1 (20)  0 (0) | 2 (22)  0 (0)  4 (44)  1 (11)  2 (22) | 0.09 | 1.00  <0.01 (<0.01, >99)  2.73 (0.88, 8.55)  1.42 (0.37, 5.44)  3.80 (0.57, 25.5) | 0.99  0.02*  0.47  0.05* |
| **Concern for Seizure**, N (%)  0: No  1: Yes  (Deceased/lost to follow-up, N=4) | 12 (86)  2 (14) | 7 (70)  3 (30) | 3 (60)  2 (40) | 1 (14)  6 (86) | 0.02* | 2.94 (1.02, 8.43) | 0.005** |

* P value significant at <.05 (not adjusted using Bonferroni correction). ** P value significant or marginally significant at <.006 (adjusted using Bonferroni correction). ^a^ P value from Fisher exact test; ^b^ P value from ordinal/multinomial logistic regression.

**eTable 6: Association of Baseline Factors with Task-fMRI.**

| **Factor** | **Task-fMRI** | | | **P value ^a^** | **Ordinal/Multinomial Logistic Regression** | |
| --- | --- | --- | --- | --- | --- | --- |
|  | **0: Normal**  **(N=30)** | **2: Moderately abnormal (N=3)** | **3: Severely abnormal (N=3)** |  | **Odds Ratio**  **(99.4% CI)** | **P value ^b^** |
| **HIE**, N (%)  0: No  1: Yes | 9 (30)  21 (70) | 1 (33)  2 (77) | 1 (33)  2 (67) | 0.99 | 0.94 (0.34, 2.62) | 0.87 |
| **HIE**, N (%)  1: Mild  2: Moderate  3: Severe | 12 (57)  6 (29)  3 (14) | 0 (0)  0 (0)  2 (100) | 0 (0)  1 (50)  1 (50) | 0.02* | 2.87 (0.72, 11.4) | 0.04* |
| **Neuro Exam**, N (%)  0: Normal  1: Mildly abnormal  2: Moderately abnormal  3: Severely abnormal | 9 (30)  7 (23)  11 (37)  3 (10) | 0 (0)  1 (33)  1 (33)  1 (33) | 0 (0)  1 (33)  1 (33)  1 (33) | 0.57 | 1.64 (0.66, 4.07) | 0.14 |
| **Consciousness (Day 0-5)**, N (%)  0: Normal  1: Irritable but arouses easily  2: Wakes up only to painful stimulation  3: Coma  4: Episodically arousable | 8 (27)  12 (40)  7 (23)  2 (7)  1 (3) | 0 (0)  1 (33)  2 (67)  0 (0)  0 (0) | 0 (0)  1 (33)  1 (33)  1 (33)  0 (0) | 0.55 | 1.67 (0.68, 4.12) | 0.12 |
| **Death**, N (%)  0: No  1: Yes | 28 (93)  2 (7) | 3 (100)  0 (0) | 2 (67)  1 (33) | 0.43 | 1.00  1.66 (0.44, 6.32) | 0.30 |
| **Discharge Condition**, N (%)  0: Normal  1: Mild support  2: Moderate deficits  3: Deceased | 15 (50)  12 (40)  1 (3)  2 (7) | 1 (33)  2 (67)  0 (0)  0 (0) | 0 (0)  1 (33)  1 (33)  1 (33) | 0.15 | 2.01 (0.77, 5.25) | 0.05* |
| **Outpatient Development**, N (%)  0: Normal  1: Mild delay  2: Moderate delay or focal finding on exam  3: Severe findings  4: Deceased | 16 (55)  1 (3)  6 (21)  4 (14)  2 (7) | 1 (33)  0 (0)  2 (67)  0 (0)  0 (0) | 0 (0)  0 (0)  1 (33)  1 (33)  1 (33) | 0.24 | 1.78 (0.72, 4.38) | 0.08 |
| **Outpatient Motor-Tone**, N (%)  0: Normal  1: Mild increased tone or weakness  2: Moderate increased tone or weakness  3: Severely increased tone or weakness  4: Deceased | 14 (48)  3 (10)  6 (21)  4 (14)  2 (7) | 2 (67)  0 (0)  1 (33)  0 (0)  0 (0) | 0 (0)  0 (0)  1 (33)  1 (33)  1 (33) | 0.45 | 1.55 (0.64, 3.73) | 0.17 |

* P value significant at <.05 (not adjusted using Bonferroni correction). ** P value significant or marginally significant at <.006 (adjusted using Bonferroni correction). ^a^ P value from Fisher exact test; ^b^ P value from ordinal/multinomial logistic regression.

**eTable 7: Association of Baseline Factors with Magnetic Resonance Spectroscopy (MRS).**

| **Factor** | **MRS** | | | | **P value ^a^** | **Ordinal/Multinomial**  **Logistic Regression** | |
| --- | --- | --- | --- | --- | --- | --- | --- |
|  | **0: Normal (N=15)** | **1: Mildly abnormal (N=11)** | **2: Moderately abnormal (N=5)** | **3: Severely abnormal (N=9)** |  | **Odds Ratio**  **(99.4% CI)** | **P value ^b^** |
| **HIE**, N (%)  0: No  1: Yes | 1 (8)  11 (92) | 0 (0)  5 (100) | 0 (0)  1 (100) | 0 (0)  3 (100) | 1.0 | **-** | **-** |
| **HIE**, N (%)  1: Mild  2: Moderate  3: Severe | 7 (64)  3 (27)  1 (9) | 4 (80)  0 (0)  1 (20) | 0 (0)  1 (100)  0 (0) | 0 (0)  0 (0)  3 (100) | 0.02* | 3.28 (0.82, 13.1) | 0.02* |
| **Neuro Exam**, N (%)  0: Normal  1: Mildly abnormal  2: Moderately abnormal  3: Severely abnormal | 4 (33)  3 (25)  4 (33)  1 (8) | 0 (0)  1 (20)  4 (80)  0 (0) | 0 (0)  1 (100)  0 (0)  0 (0) | 0 (0)  1 (33)  0 (0)  2 (67) | 0.10 | 2.21 (0.70, 6.99) | 0.06 |
| **Consciousness (Day 0-5)**, N (%)  0: Normal  1: Irritable but arouses easily  2: Wakes up only to painful stimulation  3: Coma  4: Episodically arousable | 4 (33)  6 (50)  2 (17)  0 (0)  0 (0) | 0 (0)  1 (20)  3 (60)  1 (20)  0 (0) | 0 (0)  1 (100)  0 (0)  0 (0)  0 (0) | 0 (0)  1 (33)  1 (33)  1 (33)  0 (0) | 0.20 | 2.45 (0.75, 8.02) | 0.04* |
| **Death**, N (%)  0: No  1: Yes | 12 (100)  0 (0) | 5 (100)  0 (0) | 1 (100)  0 (0) | 2 (67)  1 (33) | 0.19 | - | - |
| **Discharge Condition**, N (%)  0: Normal  1: Mild support  2: Moderate deficits  3: Deceased | 7 (58)  5 (42)  0 (0)  0 (0) | 3 (60)  2 (40)  0 (0)  0 (0) | 1 (100)  0 (0)  0 (0)  0 (0) | 1 (33)  1 (33)  0 (0)  1 (33) | 0.52 | 1.47 (0.48, 4.54) | 0.34 |
| **Outpatient Development**, N (%)  0: Normal  1: Mild delay  2: Moderate delay or focal finding on exam  3: Severe findings  4: Deceased | 8 (73)  0 (0)  1 (9)  2 (18)  0 (0) | 3 (60)  0 (0)  1 (20)  1 (20)  0 (0) | 1 (100)  0 (0)  0 (0)  0 (0)  0 (0) | 1 (33)  0 (00)  1 (33)  0 (0)  1 (33) | 0.56 | 1.00  -  1.84 (0.38, 9.05)  0.63 (0.05, 8.52)  >99 (<0.01, >99) | -  -  0.29  0.62  1.00 |
| **Outpatient Motor-Tone**, N (%)  0: Normal  1: Mild increased tone or weakness  2: Moderate increased tone or weakness  3: Severely increased tone or weakness  4: Deceased | 7 (64)  1 (9)  1 (9)  2 (18)  0 (0) | 3 (60)  0 (0)  1 (20)  1 (20)  0 (0) | 1 (100)  0 (0)  0 (0)  0 (0)  0 (0) | 1 (33)  0 (0)  1 (33)  0 (0)  1 (33) | 0.76 | 1.00  <0.01 (<0.01, >99)  1.76 (0.35, 8.74)  0.58 (0.04, 8.29)  >99 (<0.01, >99) | 1.0  0.33  0.57  1.00 |

* P value significant at <.05 (not adjusted using Bonferroni correction). ** P value significant or marginally significant at <.006 (adjusted using Bonferroni correction). ^a^ P value from Fisher exact test; ^b^ P value from ordinal/multinomial logistic regression.

**eTable 8: Association of Baseline Factors with electroencephalogram (EEG).**

| **Factor** | **EEG** | | | | **P value ^a^** | **Ordinal/Multinomial**  **Logistic Regression** | |
| --- | --- | --- | --- | --- | --- | --- | --- |
|  | **0: Normal**  **(N=3)** | **1: Mild background abnormality only**  **(N=22)** | **2: Seizure**  **(N=13)** | **3: Flat**  **(N=1)** |  | **Odds Ratio**  **(99.4% CI)** | **P value ^b^** |
| **HIE**, N (%)  0: No  1: Yes | 0 (0)  3 (100) | 7 (32)  15 (68) | 4 (31)  9 (69) | 1 (100)  0 (0) | 0.39 | 0.51 (0.11, 2.36) | 0.22 |
| **HIE**, N (%)  1: Mild  2: Moderate  3: Severe | 3 (100)  0 (0)  0 (0) | 8 (53)  5 (33)  2 (13) | 3 (33)  2 (22)  4 (44) | 0 (0)  0 (0)  0 (0) | 0.29 | 4.56 (0.66, 31.32 | 0.03* |
| **Neuro Exam**, N (%)  0: Normal  1: Mildly abnormal  2: Moderately abnormal  3: Severely abnormal | 2 (67)  0 (0)  1 (33)  0 (0) | 6 (27)  7 (32)  7 (32)  2 (9) | 2 (15)  4 (31)  5 (39)  2 (15) | 0 (0)  0 (0)  0 (0)  1 (100) | 0.55 | 2.75 (0.73, 10.3) | 0.04* |
| **Consciousness (Day 0-5)**, N (%)  0: Normal  1: Irritable but arouses easily  2: Wakes up only to painful stimulation  3: Coma  4: Episodically arousable | 2 (67)  0 (0)  1 (33)  0 (0)  0 (0) | 5 (23)  9 (41)  6 (27)  1 (5)  1 (5) | 2 (15)  6 (46)  4 (31)  1 (8)  0 (0) | 0 (0)  0 (0)  0 (0)  1 (100)  0 (0) | 0.43 | 2.13 (0.59, 7.69) | 0.11 |
| **Death**, N (%)  0: No  1: Yes | 3 (100)  0 (0) | 21 (95)  1 (5) | 12 (92)  1 (8) | 0 (0)  1 (100) | 0.14 | 1.00  7.03 (0.31, 161) | 0.09 |
| **Discharge Condition**, N (%)  0: Normal  1: Mild support  2: Moderate deficits  3: Deceased | 1 (33)  2 (67)  0 (0)  0 (0) | 12 (55)  8 (36)  1 (5)  1 (5) | 6 (46)  5 (38)  1 (8)  1 (8) | 0 (0)  0 (0)  0 (0)  1 (100) | 0.52 | 1.66 (0.44, 6.23) | 0.29 |
| **Outpatient Development**, N (%)  0: Normal  1: Mild delay  2: Moderate delay or focal finding on exam  3: Severe findings ^d^  4: Deceased | 1 (50)  0 (0)  0 (0)  1 (50)  0 (0) | 14 (64)  1 (5)  4 (18)  2 (9)  1 (5) | 3 (23)  1 (8)  6 (46)  2 (15)  1 (8) | 0 (0)  0 (0)  0 (0)  0 (0)  1 (100) | 0.09 | 3.46 (0.79, 15.2) | 0.02* |
| **Outpatient Motor-Tone**, N (%)  0: Normal  1: Mild increased tone or weakness  2: Moderate increased tone or weakness  3: Severely increased tone or weakness  4: Deceased | 1 (50)  0 (0)  0 (0)  1 (50)  0 (0) | 14 (64)  2 (9)  3 (14)  2 (9)  1 (5) | 3 (23)  0 (0)  7 (54)  2 (15)  1 (8) | 0 (0)  0 (0)  0 (0)  0 (0)  0 (0) | 0.02* | 1.00  0.65 (0.01, 38.8)  7.06 (0.75, 66.5)  1.39 (0.10, 20.2)  18.65 (0.46, 754) | 0.77  0.02*  0.73  0.03* |
| **Concern for Seizure**, N (%)  0: No  1: Yes  (Deceased/Lost to follow-up, N=4) | 2 (100)  0 (0) | 18 (86)  3 (14) | 2 (17)  10 (83) | 0 (0)  0 (0) | 0.0001** | 1.00  30.4 (2.02, 458) | 0.0005** |

* P value significant at <.05 (not adjusted using Bonferroni correction). ** P value significant or marginally significant at <.006 (adjusted using Bonferroni correction). ^a^ P value from Fisher exact test; ^b^ P value from ordinal/multinomial logistic regression.

**eTable 9. Summary of *P* values <0.05 Indicating Association of Resting State Networks and Acute Tests with Outcomes.**

|  |  | **Resting State Networks** | | | | **Acute Tests** | | | |
| --- | --- | --- | --- | --- | --- | --- | --- | --- | --- |
|  |  | **BG** | **Lang/FP** | **DMN** | **RS-SOZ** | **a-MRI** | **Task-fMRI** | **MRS** | **EEG** |
| **Acute**  **Cond.** | **HIE Severity** | 0.03^OLR^ |  | 0.03^FE^ |  | 0.002^OLR^ | 0.02^FE^ | 0.02^OLR^ | 0.03^OLR^ |
|  | **Neuro Exam** | 0.01^MLR^ | 0.03^OLR^ | 0.002^FE^ |  | ***0.0002^FE^*** |  |  | 0.04^OLR^ |
|  | **Consciousness** |  | 0.03^OLR^ | 0.05^OLR^ |  | 0.04^OLR^ |  | 0.04^OLR^ |  |
| **Outcomes** | **Death** | 0.05^LR^ | 0.008^FE^ | 0.01^FE^ |  |  |  |  |  |
|  | **Discharge Condition** | 0.02^FE^ | 0.002^OLR^ | 0.006^OLR^ |  |  | 0.05^OLR^ |  |  |
|  | **Outpatient FU Development** | ***0.0002^OLR^*** | 0.002^OLR^ | 0.02^MLR^ | 0.007^FE^ | 0.007^OLR^ |  |  | 0.02^OLR^ |
|  | **Outpatient FU Motor tone** | ***0.0003^OLR^*** | 0.01^OLR^ | 0.02^MLR^ | 0.003^OLR^ | 0.02^MLR^ |  |  | 0.02^FE^ |
|  | **Outpatient FU Seizure** |  |  |  | 0.02^OLR^ | 0.005^OLR^ |  |  | ***0.0001^FE^*** |

Abbreviations of statistical tests*:* FE=Fisher exact; LR=logistic regression; OLR=ordinal logistic regression; MLR=multinomial logistic regression. Other: BG=basal ganglia network; Lang/FP=language and/or frontoparietal network; DMN=default mode network; RS-SOZ, resting-state seizure onset zone or abnormal findings concerning for seizure related pathology; a-MRI=anatomical magnetic resonance imaging; task-fMRI=task functional magnetic resonance imaging; MRS=magnetic resonance spectroscopy; EEG=electroencephalogram; HIE=hypoxic ischemic encephalopathy; FU=follow-up.

The lowest P value was selected from either FE, LR/OLR/MLR analyses as shown in Tables S1-S8 above. **BLUE** cells indicate P value ≤ 0.006 after Bonferroni correction for multiple comparisons of each testing modality. P value < 0.0007 (***bolded and italic***) after correcting of all comparisons in Tables S1-S8. **Blacked out** cells were not analyzed due to lack of hypothesis.
